# Differential effect of corticosteroid treatment on Influenza, SARS, MERS, and SARS-CoV-2 patients: A meta-analysis and systematic review

**DOI:** 10.1101/2021.03.22.21254104

**Authors:** Sobur Ali, Salman Zahir Uddin, Md. Nayem Dewan, M Moniruzzaman, Mir Himayet Kabir, Mohammad Rafiqul Islam, Hossain Monir, Shanewaz Hossan, Tanvir Noor Nafiz, Rumana Rashid, Khan Mohammad Imran

## Abstract

**Background:** Corticosteroid has been used to manage inflammation caused by many diseases including respiratory viral infections. Many articles are available to support the good and bad side of this steroid use but remain inconclusive. To find some evidence about the safety of the drug, we investigated the effect of corticosteroids on the mortality of patients with respiratory viral infections including SARS-CoV-2, SARS, MERS, and Influenza.

**Method:** We searched articles in PubMed, Scopus, Cochrane, Medline, Google Scholar, and Web of Science records using keywords “corticosteroid” or “viral infection” or “patients” or “control study”. Mortality was the primary outcome.

**Result:** Our selected 24 studies involving 16633 patients were pooled in our meta-analysis. Corticosteroid use and overall mortality were not significantly associated (P=0.176), but in subgroup analysis, corticosteroid use was significantly associated with lower mortality in the case of SARS (P=0.003) but was not significantly associated with mortality for Influenza (H1N1) (P=0.260) and SARS-CoV-2 (P=0.554). Further analysis using study types of SARS-CoV-2, we found that corticosteroid use was not significantly associated with mortality in the case of retrospective cohort studies (P=0.256) but was significantly associated with lower mortality in the case of randomized control trials (P=0.005). Our findings uncover how the outcome of particular drug treatment for different diseases with comparable pathogenesis may not be similar and, RCTs are sometimes required for robust outcome data.

**Conclusion:** At the beginning of the COVID-19 pandemic, data of corticosteroid use from other viral infections along with COVID-19 observational and retrospective cohort studies created confusion of its effect, but randomized control trials showed that corticosteroid can be used to treat COVID-19 patients.

## 1. Introduction

### 1.1 Respiratory Virus

Respiratory infections represent the leading cause of disease and economic burden [1]. They are linked with a number of clinical symptoms from upper respiratory tract self-limited infections to more severe conditions in the lower respiratory tract, for example, pneumonia. Respiratory viruses (RVs) includes but are not limited to influenza virus, adenovirus, human coronavirus, human metapneumovirus, rhinovirus (RV), parainfluenza, and respiratory syncytial virus (RSV), may cause severe diseases such as pneumonia and bronchiolitis and/or exacerbate chronic obstructive pulmonary disease (COPD) and asthma. Moreover, the Avian influenza virus (H5N1), SARS coronavirus (SARS-CoV), and MARS coronavirus (MARS-CoV) have emerged in recent years as potential threats to the public health attracting global attention. The coronavirus disease 2019 (COVID-19) pandemic caused by severe acute respiratory syndrome coronavirus 2 (SARS-CoV-2) originating in Wuhan, China, is swiftly and continuously spreading globally and responsible for significant respiratory morbidity and mortality [2].

### 1.2 Corticosteroid

Corticosteroids, often known as steroids, are classes of hormones secreted by the adrenal cortex, including glucocorticoids (GCs) and mineralocorticoids [3]. Nevertheless, the term “corticosteroids” is commonly used to denote glucocorticoids. Corticosteroids control various cellular functions such as metabolism, development, cognition, inflammation, and homeostasis [4]. Corticosteroids are being used regularly in clinics to treat autoimmune and inflammatory pathologies such as allergy, septic shock, asthma, rheumatoid arthritis, multiple sclerosis, and inflammatory bowel disease. The adverse effects of prolonged glucocorticoid therapy are well established and exceedingly common which limits the therapeutic benefits. Some side effects of certain medical conditions after taking corticosteroid are glucose intolerance and diabetes, osteoporosis and fracture, muscle wasting, central obesity, growth retardation, depression, hypertension cataracts, and increased risk of viral and bacterial infections [5].

To the current day, nevertheless, corticosteroids continue to be the backbone in the treatment of autoimmune disorders and inflammatory diseases, and they are prescribed as an immunosuppressant after organ transplantation and as lymphocytic in chemotherapeutic treatments [6].

### 1.3 Corticosteroid and immunosuppression

Corticosteroids, especially glucocorticosteroids, have inhibitory properties on a wide range of immune responses [7]. They are steroid hormones with extensive effects. They often exert anti-inflammatory and potent immune-suppressive effects when pharmacologically driven. These are typically used to prevent the harmful effects of the inflammatory cascades in severe infections [8] and that’s why they are used to treat inflammatory diseases due to the inhibition of B cells and T cells mediated immune responses [7]. To date, the immunomodulatory function of glucocorticoid analogs has been difficult to discern from their untoward effects, which reflects the fact that most of the activities of cortisol are regulated by a particular molecule of the nuclear receptors. For the future production of more active immunosuppressive agents, a better comprehension of the molecular base of steroids on the immune response is thus necessary [9].

While glucocorticoids play a role in stimulating lymphocyte apoptosis and in altering leukocyte movement and redistribution, the key component of their action is the suppression of cytokine gene expression, resulting in a reduced release of interferons (IFN-α), interleukins (IL-2, IL-6), and tumor necrosis factor (TNF-α) [9].

### 1.4 Use of corticosteroid in respiratory viral infection

Even after controversy, it is a very common trend to administer corticosteroids to influenza patients, especially pandemic influenza virus. The use of corticosteroids increases the chance of infection by various microorganisms and it is significantly associated with mortality (OR 1.98, 95% CI 1.62-2.43, p < 0.00001) and nosocomial infection (OR 3.16, 95% CI 2.09-4.78, p < 0.00001) [10]. Patients with pandemic H1N1 viral infection whose rapidly deteriorating pneumonia leads to acute lung injury (ALI) or acute respiratory distress syndrome (ARDS) along with several organ dysfunctions were found associated with an increased rate of mortality (14–41%) [11]. In clinical practice, the application of adjuvant corticosteroid treatment of those patients is very common. Other studies have shown that systemic corticosteroid therapy may inhibit the inflammatory cascade reaction in patients with community-acquired pneumonia [12]. It has also been reported that corticosteroid use was not significantly associated with 90-day mortality (adjusted odds ratio, 0.75; 95% confidence interval, 0.52-1.07; P=0.12) but found to be associated with the delay in RNA clearance of MERS coronavirus (adjusted hazard ratio, 0.35; 95% CI, 0.17-0.72; P=0.005) [13].

### 1.5 Covid-19 and corticosteroid

To date, 115 million people are infected by the SARS-CoV-2 virus worldwide and caused 2.5 million death [14]. Although vaccination has been started, only 269 million out of 7.79 billion have been given a single shot and 56 million people have been fully vaccinated (0.72% of the total population) [15].

The complex condition of severely ailing patients with COVID-19 leads to an array of protocols employing complementary treatments including the use of corticosteroids for the treatment of phase IIb-III COVID-19 patients in hospital[16–18]. The vital point is that corticosteroids might be useful in checking cytokine and chemokine storm-mediated alveolar/pulmonary damage. In this condition, corticosteroids are used as immunosuppressants (inhibition of cytokine production) resulting in impaired lymphocyte proliferation and delay in the clearance of the virus [17,19]. Nevertheless, there are several studies reported the use of corticosteroids for the management of ARDS caused by several severe coronavirus infections such as severe acute respiratory syndrome (SARS) [20] and the Middle East respiratory syndrome (MERS) [13] where diffused alveolar damage and histological pulmonary inflammation are common [21].

Patients with COVID-19 are frequently treated with corticosteroids regardless of the deficiency of effectiveness evidence from clinical studies [22]. Moreover, the present temporary direction from WHO on clinical management of COVID-19 patients recommends against corticosteroid use if not prescribed for another reason. A study conducted at Wuhan Union Hospital retrospectively reviewed forty-six severe COVID-19 pneumonia hospitalized patients for the use of intravenous methylprednisolone (dose 1-2mg/kg/d for 5-7 days in 26 patients) in 26 of them and the other 20 as control. Data of this study indicate that early, low-dose, and short-term use of corticosteroids in patients with severe COVID-19 was correlated with the quick improvement of various symptoms and absorption of lung focus [23]. In another study, COVID-19 patients treated with a corticosteroid had adverse clinical symptoms, further aberrations on chest CT, and an elevated inflammation index. Their data indicates that corticosteroid use could not stimulate virus clearance time, length of hospital stay, or overall duration of symptoms persistence in mild COVID-19 patients [24]. Studies have also revealed that corticosteroid treatment in patients with SARS-CoV, MERS-CoV, and SARS-CoV-2, infections had delayed virus clearance and did not significantly improve survival rate, decrease hospitalization time or rate of ICU admission and/or requirement of mechanical ventilation [25].

To make evidence-based recommendations for clinicians about the use of corticosteroids in COVID-19 patients require systematic summaries and meta-analysis of the available evidence. Therefore, the contentious results of corticosteroid use in SARS, MERS, influenza pneumonia patients lead us to conduct a systematic review and meta-analysis of all observational, retrospective cohort, and randomized control studies that have compared mortality, secondary infection, ICU stays, and hospital stay of patients who received corticosteroid treatment with the patients who did not. We aimed to recognize the safety and efficacy of corticosteroids as well as their role in clinical outcomes in SARS, MARS, influenza, and COVID-19 patients and also aimed to find out differences in the outcome of different types of studies such as retrospective cohort studies (RCS) and randomized control trials (RCT).

## 2. Methods

### 2.1 Articles search strategies

A literature search was performed in PubMed, Scopus, Cochrane Central Register of Controlled Trials (CENTRAL), Medline, Google Scholar, and Web of Science records using keywords such as: “corticosteroid” or “viral infection” or “patients” or “control study”. No limitation on language, publication type, and location of the word in the article (title, abstract and main text) was used. Our search consisted of articles from 1980 to September 2020. References from similar review articles and selected articles were also checked for matching articles with our selection criteria. We have used PRISMA 2009 checklist for systematic reviews and meta-analysis (www.prisma-statement.org) [26] and available in supplementary data (Suppl. Table ST1).

### 2.2 Criteria for inclusion and exclusion of articles

Articles were selected for this meta-analysis based on the following inclusion criteria i) patients with respiratory viral infections in each study were treated with corticosteroid ii) study had corticosteroid treatment group and control group (placebo-treated or untreated/matched) iii) study had primary outcome data on mortality, and any of the secondary outcomes such as secondary infection rate, length of ICU stay, length of hospital stay, duration of mechanical ventilation, and number patients in mechanical ventilation. Studies on non-respiratory viral infections, non-English articles, reviews, case study and articles that didn’t have full texts were excluded from this analysis.

### 2.3 Article selection

Every search was grouped and assigned to all the authors to primarily select articles. Next, those selected articles were reassigned to different authors to make a final selection based on our inclusion and exclusion criteria. Titles of selected articles from all of the authors were then copy-pasted into a shared google sheet that was conditionally formatted for matching to avoid duplications. The corresponding author then checked all the selected articles again to confirm inclusion and exclusion criteria match.

### 2.4 Data extraction

Selected articles were grouped and assigned to all the authors to extract data and put into a google sheet that only the corresponding author had access to. Next, those groups of articles were reassigned to a different author to extract data and record in a separate google sheet. Two sheets were then matched, and any discrepancies were resolved by group meetings via Zoom. Authors were asked to extract information on authors, title, year of publication, number of patient in the treatment arm, number of patient in the control arm, a corticosteroid used, placebo-control or not, type of study, the mortality rate (control/corticosteroid), secondary infection rate (bacterial), secondary infection rate (viral), male/female (% of the total patient), male/female (% of control/corticosteroid group), the mean age of the total patient, mean age of the dead patient, length of hospital stay, length of ICU stay, dead patients underlying health conditions, comorbidities of patients, name of viral infection, length of mechanical ventilation, Viral/RNA clearance time/viral shedding duration, and the median daily dosage of corticosteroid. For any unavailable data in the article during extraction, authors from that article were contacted by emails to see if those data were available.

### 2.5 Quality assessment

The risk of bias quality of each of the selected studies was individually evaluated by two authors according to the Newcastle-Ottawa Scale for comparative observational studies and nonrandomized trials and the Cochrane Risk of Bias tool for RCTs [27,28]. Disputes related to quality at any point were settled through zoom meetings with the other authors until the agreement was achieved.

### 2.6 Statistical Analysis

All statistical analysis was performed by using statistical software STATA 13 [29]. The hypothesis was tested using the Mann-Whitney U test. Z statistic and P-value <0.05 were used to test the significance. All the results of the meta-analysis are displayed in the forest plot. Frequencies and proportions are used to report binary variables. On the other hand, mean and standard deviation are used to report continuous variables. Chi-square statistics are used to test statistical heterogeneity. A random-effects model meta-analysis is performed under the appearance of statistical heterogeneity. The derSimonian-Laird estimator is used to estimating between-study variance (τ^2^) in a random-effects model. For binary data, we have calculated risk ratio (RR) as a relative effect along with a 95% confidence interval (CI). On the other hand, the standard mean difference (SMD) is calculated as a relative effect for continuous data along with 95% CI. We have conducted subgroup analysis based on different types of viruses. A funnel plot is used to check the publication bias and Egger’s test of the intercept is used to quantify the asymmetry of the funnel plot and to perform a statistical test.

## 3. Results

Our search initially identified 2365 records, of which 1344 records were from PubMed, 450 from Cochrane, 101 from Medline, 241 from Scopus, 130 from Web of science, and 100 (first ten pages) from Google scholar. After screening and reading through the title, abstract, and main text, we discarded 1835 studies due to not fulfilling our selection criteria. Twenty-four studies finally satisfied our exclusion and inclusion criteria and were used in this analysis. Further details of the exclusion and inclusion of articles are shown in figure 1.

**Figure 1.**
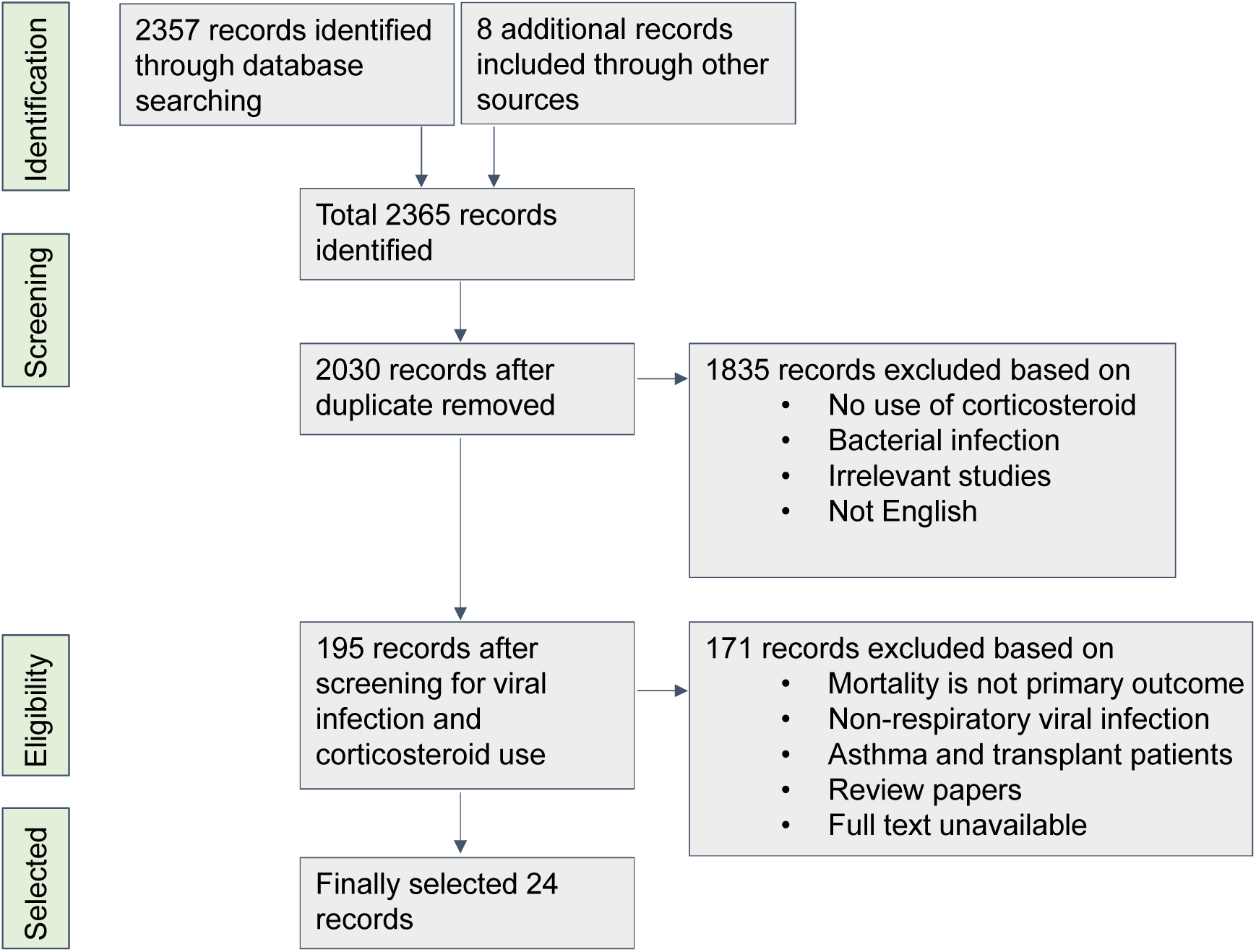
Schematic diagram of the article selection process.

### 3.1 Description of the selected studies

All 24 of our selected articles compared outcomes of corticosteroid with the control group and our primary outcome mortality was recorded in all 24 studies [13,30,39–48,31,49–52,32–38]. Length of hospital stay (days) was analyzed in 3 studies [13,48,50], secondary infection rate was recorded in 12 studies [32,37,50,51,42–49], length of mechanical ventilation was recorded in 5 studies [39,42,49–51], number of patients in mechanical ventilation (MV) was presented in 14 studies [13,31,48,50,51,33,34,38,41,42,45–47], length of ICU stay (days) was reported in 6 studies [13,43,48–51]. Twenty-three articles included patients with the respiratory viral infection and one article included patients with acute respiratory distress syndrome. Of the 23 respiratory viral infection articles, eight were H1N1, one H7N9, one influenza, one SARS, one MERS, and eleven SARS-CoV-2. Detailed characteristics of studies can be found in supplementary table ST1. We have included studies published between 2007 and September 2020. The outcome between corticosteroid and non-corticosteroid of all 24 studies were compared based on mortality. We have included 16633 patients into our final meta-analysis and systematic review from all 24 studies. Among all the patients, about 7634 patients were treated with corticosteroid and the remaining 8999 patients were in the control group. Baseline characteristics of patients in this analysis are shown in supplementary table ST1.

### 3.2 Statistical heterogeneity

We have found significant heterogeneity in our meta-analysis of effect of corticosteroid on mortality (I^2^=95.7%, P<0.0001), secondary infection (I^2^=90.6%, P<0.0001), MV days (I^2^=70.7%, P=0.009), MV patients (I^2^=96.4%, P<0.0001), ICU days (I^2^=91.2%, P<0.0001), hospital days (I^2^=80.3%, P=0.006).

### 3.3 Overall Mortality

First, we combined all the studies and analyzed them as a whole although it is obvious that these viral diseases are different in their pathology and may not react the same but just to understand from an overall point of view. We found that the mortality rate for the patients who received corticosteroids was not significantly higher than that of patients who did not receive corticosteroids (RR: 1.23 (CI: 0.91, 1.65), Z = 1.35, P = 0.176)) (Figure 2). Next, we analyzed them based on viral types and we found that subgroup analysis based on viral types shows differential results (Figure 3). Patients with MERS–CoV, H7N9 viral types favored control over corticosteroid (Z=3.03, P=0.002 and Z= 3.05, P=0.002 respectively) and SARS study favored corticosteroid but had just one study, so the results were not very robust. Similar to the overall mortality, H1N1 and SARS-CoV-2 studies showed that the mortality of patients who received corticosteroids was not significantly higher than that of patients who did not receive corticosteroids (Figure 3). On the flip side, different types of SARS–CoV-2 studies showed different outcomes. Retrospective cohort studies leaned towards control over corticosteroids but did not show statistically significant differences in mortality (Z=1.14, P=0.256), whereas RCT (Z=2.84, P=0.005) and retrospective controlled cohort (Z=2.16, P=0.031) studies showed significantly lower mortality when corticosteroids were used (Figure 4).

**Figure 2.**
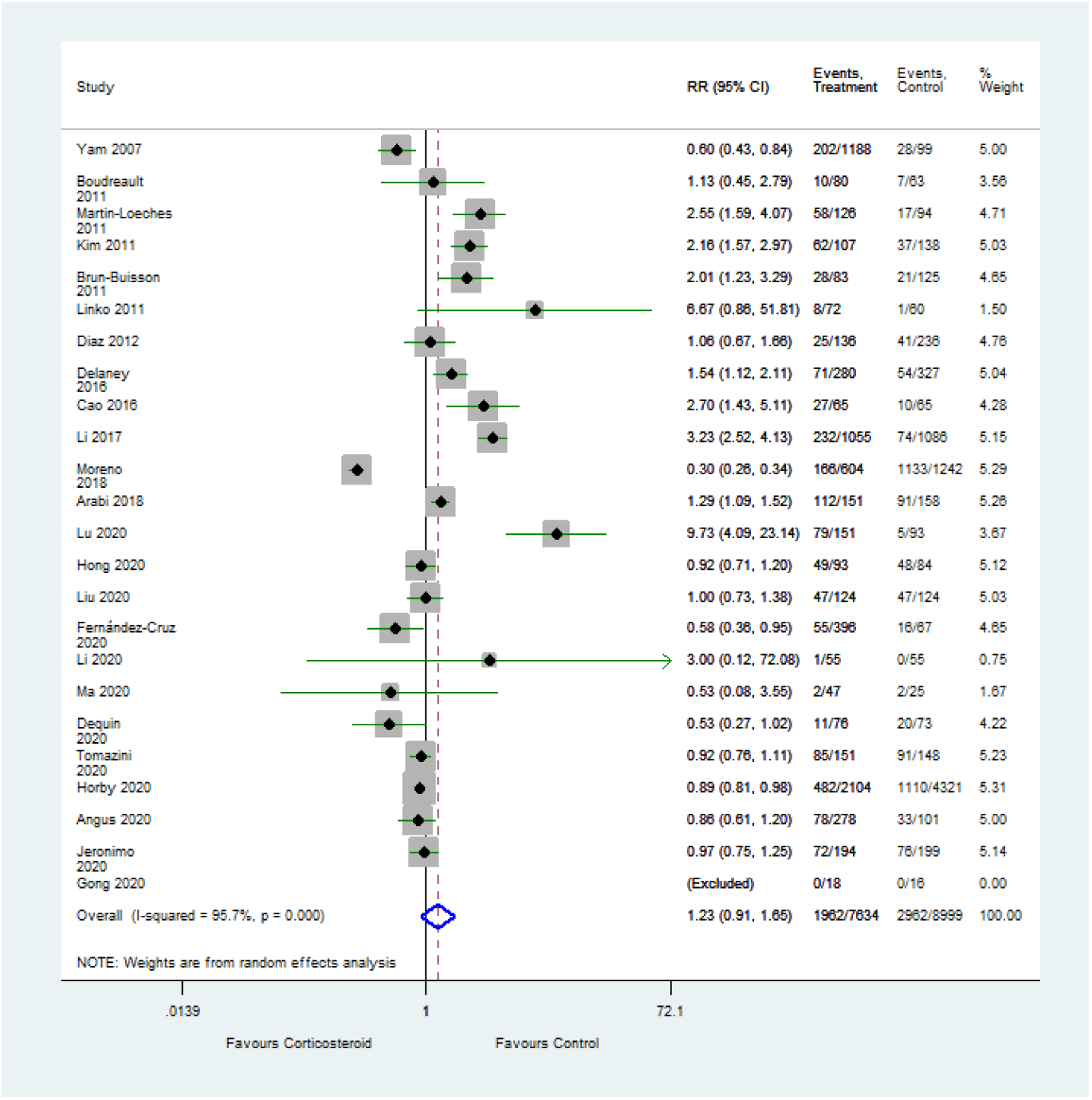
Effect of corticosteroid on mortality (Z=1.35, P=0.176, RR: Risk Ratio, horizontal line express 95% CI, Diamond represents overall estimate from the meta-analysis, squares represent effect size for each study).

**Figure 3.**
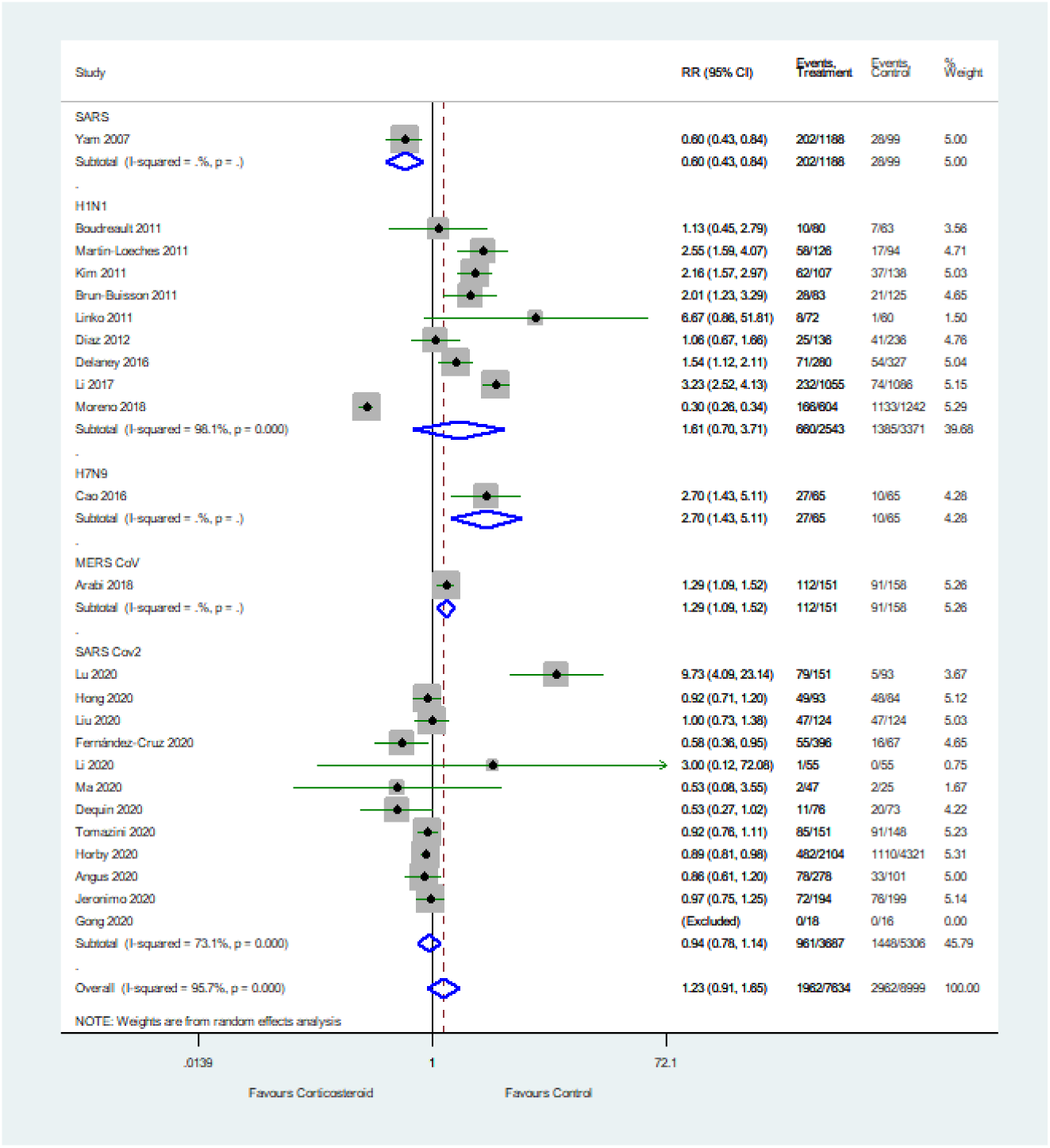
Subgroup analysis of the effect of corticosteroid on mortality of patients with SARS, H1N1, H7N9, MERS-Cov, and SARS-CoV-2 infection. RR: Risk Ratio, horizontal line express 95% CI, Diamond represents overall estimate from the meta-analysis, squares represent effect size for each study.

**Figure 4.**
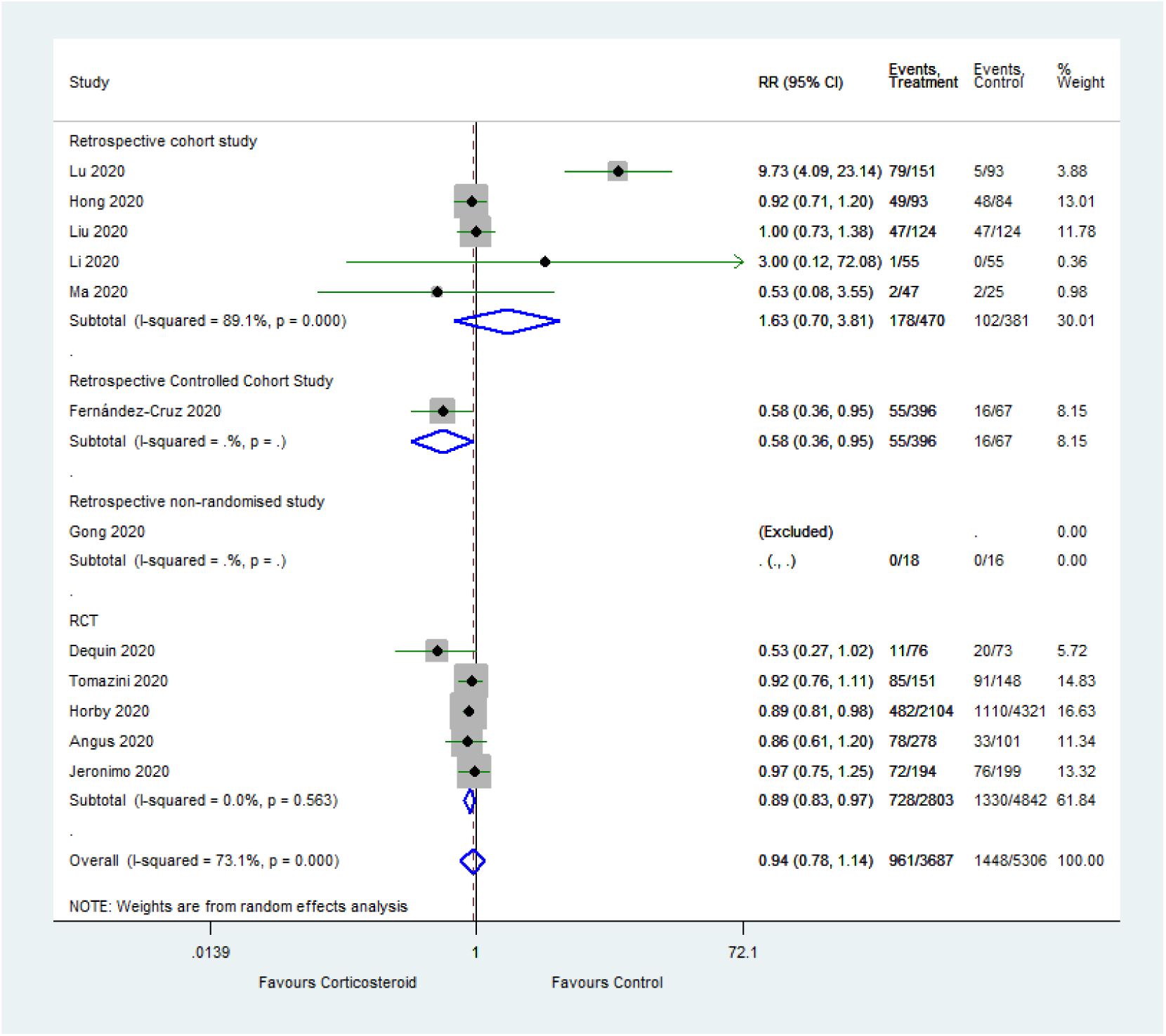
Subgroup analysis based on study type of SARS-CoV-2. RCT: Randomized control trial, RR: Risk Ratio, horizontal line express 95% CI, Diamond represents overall estimate from the meta-analysis, squares represent effect size for each study.

### 3.4 Secondary infection

Next, we analyzed if immunosuppressive corticosteroid treatment increases secondary infection rates. Secondary infection, length of hospital stays, and mortality are closely related. On the one hand, secondary infection can increase mortality and length of hospital and ICU stay; on the other hand, the length of hospital stay can increase secondary infection chances. We found that the secondary infection rate for the patients who received corticosteroid is significantly higher than the patients who did not receive corticosteroid (RR: 1.55 (CI: 1.05, 2.28), Z=2.20, P=0.028)) as shown in Figure 5. Similar results were also obtained from a subgroup analysis of patients with H1N1 viral type (RR: 1.58 (CI: 1.01, 2.46), Z=2.0, P=0.045)). Patients with SARS and H7N9 viral types found no significant corticosteroid effect on the secondary infection (Figure 5). Two SARS-CoV-2 studies included data on secondary infection, one of them is RCT [32] and the other is RCS [37]. Interestingly, similar to the mortality data RCT favored corticosteroid but RCS favored control (Figure 5).

**Figure 5.**
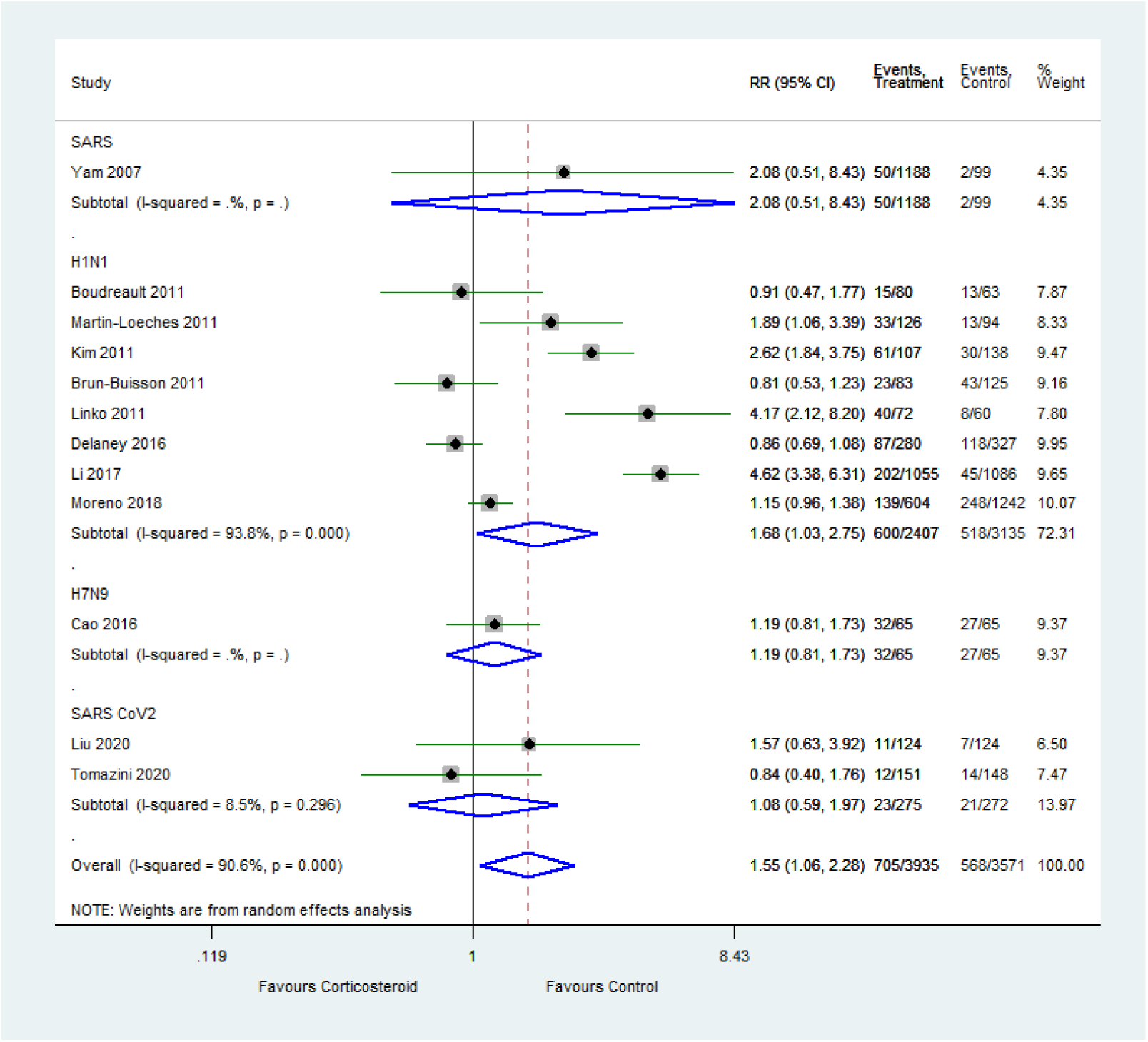
Subgroup analysis of the effect of corticosteroid on the rate of secondary infection for patients with SARS, H1N1, H7N9, and SARS-CoV-2 infection. (RR: Risk Ratio, horizontal line express 95% CI, Diamond represents overall estimate from the meta-analysis, squares represent effect size for each study).

### 3.5 Number of patients in mechanical ventilation

We were intrigued to find out how many patients were in mechanical ventilation (MV) in the corticosteroid treatment group. We found that the number of patients in mechanical ventilation was significantly higher in the corticosteroid group (RR: 1.59 (CI: 1.27, 1.99), Z=4.10, P < 0.0001)) as shown in figure 6. Subgroup analysis of patients with H1N1 (Z=2.39, p=0.017), SARS-CoV-2 (Z=2.43. p=0.015), and MERS-CoV (Z=4.04, p<0.0001) also showed us a similar effect of corticosteroid (Figure 6), but patients with H7N9 infection showed no significance (p=0.058).

**Figure 6.**
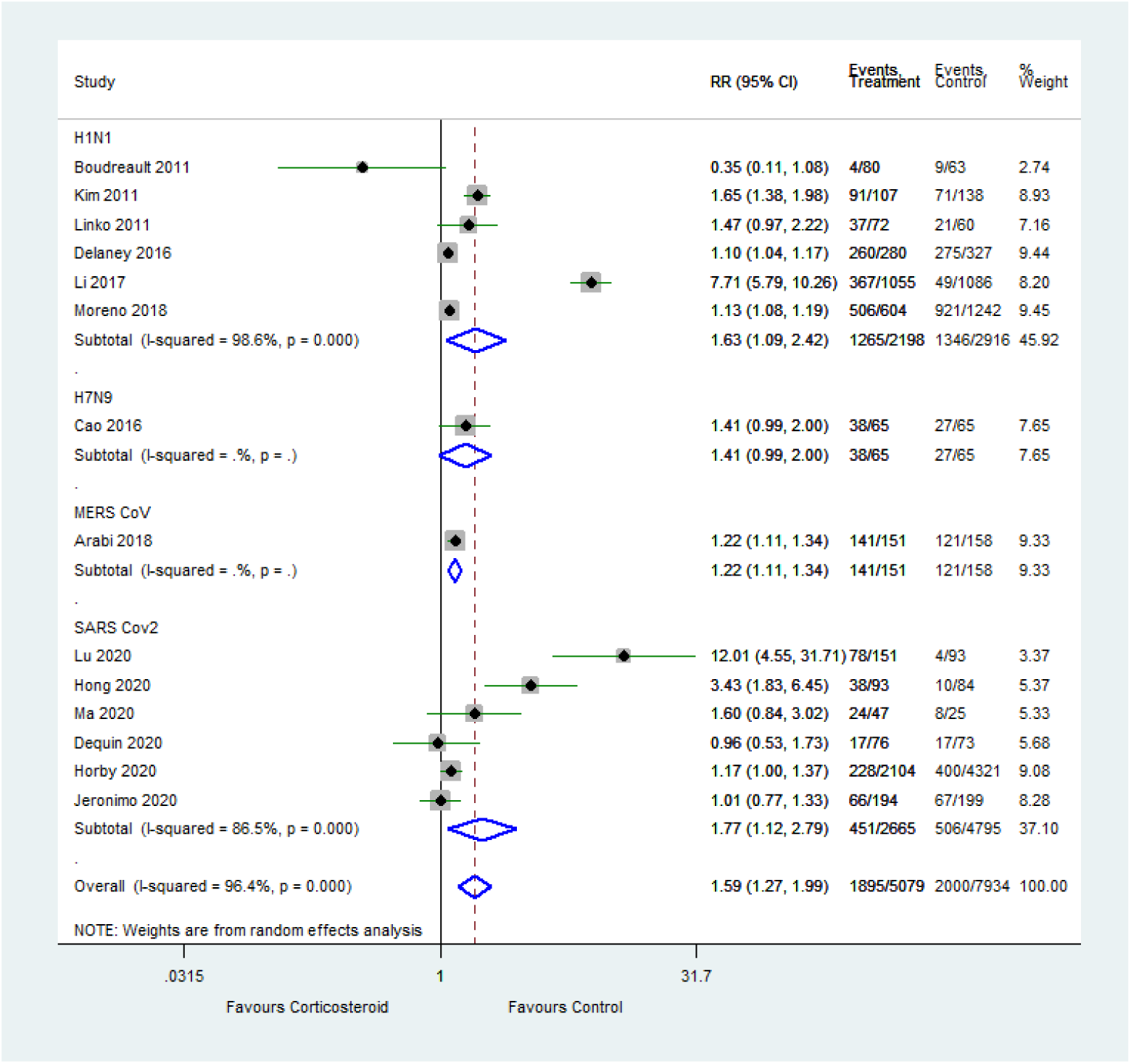
Subgroup analysis of the effect of corticosteroid on the number of patients receiving Mechanical Ventilation for H1N1, H7N9, MERS-Cov, and SARS-CoV-2 viral infections (RR: Risk Ratio, horizontal line express 95% CI, Diamond represent overall estimate from the meta-analysis, squares represent effect size for each study).

### 3.6 Length of mechanical ventilation, hospital stay, and ICU stay

If corticosteroid use is associated with prolonged stay in a hospital and a higher number of patients in mechanical ventilation, it might affect the length of mechanical ventilation requirement also. As shown in Suppl. figure S1, our analysis has revealed that the length of mechanical ventilation (days) was significantly higher in patients who received corticosteroid (SMD: 0.18 (CI: 0.02, 0.33), Z=2.26, P=0.024)). All patients were affected by the H1N1 virus.

We then analyzed three studies that had data for hospital stays to see if corticosteroid use increases patients’ hospital stay. As shown in Suppl. figure S2, length of hospital stay (days) was significantly higher for the corticosteroid group (SMD: 0.58 (CI: 0.22, 0.94), Z=3.16, P=0.002)). Subgroup analysis of patients with MERS-CoV and H1N1 showed similar results (Suppl. figure S3).

Next, we analyzed the length of ICU stay of patients receiving corticosteroids. We found that the length of ICU stay in days was significantly higher for patients who received corticosteroids (SMD: 0.48 (CI: 0.17, 0.79), Z=3.07, P=0.002). (Suppl. figure S4). Similar results were observed from a subgroup analysis of patients with H1N1 and MERS-CoV viral infections (Suppl. figure S5).

### 3.7 Risk of bias assessment

We have used funnel plot analysis of selected articles to check publication bias and found that the P-value of Egger’s test is not significant (P > 0.09), which suggests that there is no evidence of publication bias (Suppl. figure S6). In our selected studies risk of selection bias was unavoidable for the studies that were not randomized control trials. The risk of bias identified in the 24 included studies is shown in (Suppl. Table ST2 and Suppl. Table ST3). NOS scores of our selected retrospective cohort and observational studies varied from 6 to 9, suggesting that the quality of our selected studies was high [28]. Nevertheless, most selected studies had considerable comparability bias because we could not adjust for disease severity, and patients with higher disease severity have a tendency to receive corticosteroid treatment.

## 4. Discussion

This meta-analysis and systematic review included 19 cohort studies and 5 RCTs, with low to high risk of bias, which addressed the association between corticosteroid use and mortality as primary outcome and length of hospital stay, length of ICU stay, length of mechanical ventilation, number of patients required mechanical ventilation, and rate of secondary infection, as a secondary outcome with SRAS-CoV-2, SARS, MERS, and Influenza. The available data suggested that corticosteroid therapy was not significantly associated with lower mortality rate when compared using different virus types and study types but was associated with lower mortality rate when low bias, gold-standard RCTs were analyzed.

The use of corticosteroids suppresses systemic inflammation [8]. Following infection with Influenza virus, SARS-CoV, SARS-CoV-2, and MERS-CoV acute respiratory distress and acute lung injury are characterized by uncontrolled local and systemic inflammation [19,53–55]. The systemic injury is caused by an extreme host innate response with inflated migration of neutrophils, macrophages, and pro-inflammatory cytokines, promoting typical exudative edema and fibrosis which lead to diffuse alveolar damage, acute necrotizing bronchiolitis with mostly neutrophilic inflammation, and severe alveolar hemorrhage [56,57]. During these diseases, the clinical administration of corticosteroids inhibits immune reactions by suppressing inflammatory responses, inhibiting the migration of inflammatory cells from the systemic circulation to tissues by the suppression of the production of pro-inflammatory cytokines and chemokines, reducing leukocyte trafficking, and preventing immune reactions mediated by T-lymphocytes and B-lymphocytes [48,49,56,58–60]. Immune suppression occurs by the administration of corticosteroid before the inflammatory reaction reaches to deleterious position might even delay viral clearance, lead to extended viremia, and delayed viral RNA clearance, ultimately leading to an increased rate of mortality [61,62]. By applying marginal structural Cox proportional hazards modeling, one of our included studies showed that corticosteroid treatment was correlated with a substantial delay in MERS-CoV RNA clearance (aHR, 0.35; 95% CI, 0.17–0.72; P=0.005) [13] while another study has shown corticosteroids use in patients were associated with lower levels of procalcitonin (0.5 vs 0.7 ng/mL, P=0.02) [51]. On the other hand, the inflammatory response against some respiratory viral infection reaches to point where inflammation-mediated collateral damage becomes more detrimental to the host than the harm caused by the virus itself. Observational and cohort studies prior to the COVID-19 pandemic found a negative effect of corticosteroid use as we have also shown in this meta-analysis. After the pandemic, clinicians stated to use corticosteroid as an immunosuppressive drug to mitigate the cytokine storm but again most of the cohort studies showed higher mortality in the corticosteroid use. Then, randomized control trials were implemented and showed a beneficial effect of corticosteroid use. Because results from RCTs are robust that the cohort studies, it is now evident that cohorts’ studies had patient selection and other biases. In this analysis we clearly show how outcome of a drug treatment may vary depending on different diseases/pathogeneses and even on the study types within same disease/pathogenesis.

In this meta-analysis, we have found that most of the studies including one RCT [37] (except one RCT [32]) that had data on secondary infection showed that the corticosteroids group was prone to develop secondary infection probably due to the immunosuppressive effect of the drug. One of our included studies revealed that the subjects, who received corticosteroids, had an elevated rate of immunosuppression (46% vs. 32%%, P=0.03) led to higher secondary bacterial pneumonia cases (57% vs. 22%, P=<0.001) [50]. Besides, the increased rate of secondary infection due to prolonged ICU stay has also been revealed by another study [63]. Then Again, corticosteroids immune suppression might lead to developing critical illness [64]. One of our included studies has shown that the rate of shock was 43% vs 30% in the corticosteroid group vs the control group [50]. Moreover, corticosteroids treatment increased the need for invasive mechanical ventilation rate to 85% from 51% in the control group [50]. Analysis from the studies before the COVID-19 pandemic showed significant higher secondary infection for the corticosteroid group (Z=2.2, p=0.028) but two RCT of COVID-19 studies had differential data and could not generate any significance between treatment and control group suggests that more and more RCT studies are needed to reach a conclusion.

Moreover, studies before the COVID-19 pandemic (data from SARS, MERS, H1N1) showed that patients treated with corticosteroids had a significantly higher hospital stay, ICU stay, and length of mechanical ventilation. In addition, these patients were more vulnerable to superinfection, such as secondary bacterial pneumonia (57% vs. 22%, P=<0.001) or invasive pulmonary fungal infection, and aggravation of underlying conditions, which led them to stay in ICU for longer periods than the control (no-corticosteroid) group [50]. Furthermore, another study exhibited that the clinical administration of corticosteroids deferred the initiation of blocking the viral neuraminidases by neuraminidase inhibitors, with longer ICU stay in patients who did not receive neuraminidase inhibitors within 5 days of illness [65]. None of the RCTs of SARS-CoV-2 had data on hospital stay, ICU stay, and length of mechanical ventilation and due to the selection bias of cohort studies it is now apparent that RCTs are required to get a robust conclusion on these parameters.

Regardless of these outcomes, our study has some limitations. First, the strength of the conclusion made by the studies before the COVID-19 pandemic is not so strong because there was only one RCT study in our analysis, all studies that we include in this review are observational studies having many lurking variables. Now that RCTs showed a beneficial effect of corticosteroid, the deleterious effect of it from the cohort studies becomes questionable. Second, we could not separate patients who obtained corticosteroids for underlying disease (e.g., COPD). We also could not perform subgroup analysis based on doses of steroid received, early or late steroid use, or other factors. Data on dose, duration, timing, types, and rationales of corticosteroid administration and the timing and duration of antiviral therapy were very insufficient. Before starting this review one of the goals was to see what happens if corticosteroid is applied after cytokine storm vs before cytokine storm but due to data limitation, it was not possible to analyze and opens a new avenue to investigate in the future. Third, the baseline criteria of the patients can control outcomes and differences among the studies included in our analysis. For example, an association had been observed between fewer secondary infections and younger age and fewer underlying diseases.

## Conclusion

Current meta-analysis supports the use of corticosteroids to treat critically ill patients with COVID-19, because of the strength of the RCTs that shows that the administration of systemic corticosteroids was associated with lower all-cause mortality compared with usual care or placebo.

Our analysis also points out that outcome of particular drug treatments for different diseases with comparable pathogenesis may not be similar and for robust outcome data, RCTs are sometimes required.

## Data Availability

The data will be made available upon request.

## Abbreviations

RCT: Randomized Control Trial
RCS: Retrospective Cohort Study
SARS: Severe acute respiratory syndrome
MERS: Middle East Respiratory Syndrome
SARS-CoV-2: Severe Acute Respiratory Syndrome Coronavirus 2
H1N1: Hemagglutinin (H) and Neuraminidase (N)

## Supplementary Material

Supplementary material is available for this article online.

## Data availability

The data will be made available upon request.

## Ethical statement

This work did not require ethical approval as it does not involve any human or animal experiment.

