## Supplemental data for "Differential effect of corticosteroid treatment on Influenza, SARS, MERS, and SARS-CoV-2 patients: A meta-analysis and systematic review"

**Suppl table ST1**. PRISMA Checklist.

| Section/topic | # | Checklist item | Reported on page # |
|---|---|---|---|
| **TITLE** | | | |
| Title | 1 | Identify the report as a systematic review, meta-analysis, or both. | 1 |
| **ABSTRACT** | | | |
| Structured summary | 2 | Provide a structured summary including, as applicable: background; objectives; data sources; study eligibility criteria, participants, and interventions; study appraisal and synthesis methods; results; limitations; conclusions and implications of key findings; systematic review registration number. | 2-3 |
| **INTRODUCTION** | | | |
| Rationale | 3 | Describe the rationale for the review in the context of what is already known. | 4-7 |
| Objectives | 4 | Provide an explicit statement of questions being addressed with reference to participants, interventions, comparisons, outcomes, and study design (PICOS). | 7 |

| **METHODS** | | | |
|---|---|---|---|
| Protocol and registration | 5 | Indicate if a review protocol exists, if and where it can be accessed (e.g., Web address), and, if available, provide registration information including registration number. | |
| Eligibility criteria | 6 | Specify study characteristics (e.g., PICOS, length of follow-up) and report characteristics (e.g., years considered, language, publication status) used as criteria for eligibility, giving rationale. | 8 |
| Information sources | 7 | Describe all information sources (e.g., databases with dates of coverage, contact with study authors to identify additional studies) in the search and date last searched. | 8 |
| Search | 8 | Present full electronic search strategy for at least one database, including any limits used, such that it could be repeated. | 8 |
| Study selection | 9 | State the process for selecting studies (i.e., screening, eligibility, included in systematic review, and, if applicable, included in the meta-analysis). | 8-9 |
| Data collection process | 10 | Describe method of data extraction from reports (e.g., piloted forms, independently, in duplicate) and any processes for obtaining and confirming data from investigators. | 9-10 |
| Data items | 11 | List and define all variables for which data were sought (e.g., PICOS, funding sources) and any assumptions and simplifications made. | 9-10 |
| Risk of bias in individual studies | 12 | Describe methods used for assessing risk of bias of individual studies (including specification of whether this was done at the study or outcome level), and how this information is to be used in any data synthesis. | 10 |
| Summary measures | 13 | State the principal summary measures (e.g., risk ratio, difference in means). | 10 |
| Synthesis of results | 14 | Describe the methods of handling data and combining results of studies, if done, including measures of consistency (e.g., $I^2$) for each meta-analysis. | 10 |
| Risk of bias across studies | 15 | Specify any assessment of risk of bias that may affect the cumulative evidence (e.g., **publication bias**, selective reporting within studies). | 10 |

| | | | |
|---|---|---|---|
| Additional analyses | 16 | Describe methods of additional analyses (e.g., sensitivity or subgroup analyses, meta-regression), if done, indicating which were pre-specified. | 10 |
| **RESULTS** | | | |
| Study selection | 17 | Give numbers of studies screened, assessed for eligibility, and included in the review, with reasons for exclusions at each stage, ideally with a flow diagram. | 11 |
| Study characteristics | 18 | For each study, present characteristics for which data were extracted (e.g., study size, PICOS, follow-up period) and provide the citations. | 11 |
| Risk of bias within studies | 19 | Present data on risk of bias of each study and, if available, any outcome level assessment (see item 12). | 14 |
| Results of individual studies | 20 | For all outcomes considered (benefits or harms), present, for each study: (a) simple summary data for each intervention group (b) effect estimates and confidence intervals, ideally with a forest plot. | 12-13 |
| Synthesis of results | 21 | Present results of each meta-analysis done, including confidence intervals and measures of consistency. | 12-13 |
| Risk of bias across studies | 22 | Present results of any assessment of risk of bias across studies (see Item 15). | 14 |
| Additional analysis | 23 | Give results of additional analyses, if done (e.g., sensitivity or subgroup analyses, meta-regression [see Item 16]). | 12-13 |
| **DISCUSSION** | | | |
| Summary of evidence | 24 | Summarize the main findings including the strength of evidence for each main outcome; consider their relevance to key groups (e.g., healthcare providers, users, and policy makers). | 15-19 |
| Limitations | 25 | Discuss limitations at study and outcome level (e.g., risk of bias), and at review-level (e.g., incomplete retrieval of identified research, reporting bias). | 17-18 |

| | | | |
|---|---|---|---|
| Conclusions | 26 | Provide a general interpretation of the results in the context of other evidence, and implications for future research. | 18 |
| **FUNDING** | | | |
| Funding | 27 | Describe sources of funding for the systematic review and other support (e.g., supply of data); role of funders for the systematic review. | 19 |

**Suppl table ST2**. Characteristics of study and patients.

| | Author | Title | Year | Type of study | Type of Virus | Corticosteroid used | Patient in treatment arm/ control arm | Mortality rate (treated/control) | Secondary infection rate (Bacterial) (Treated/control) | Secondary infection rate (Viral) (Treated/ control) | M/F % of total patient | M/F% of Treated/control | Mean age of total patient/dead patient | length of hospital stay (treated/control) | length of ICU stay (treated/control) days | Number (%) of patient required Mechanical ventilation (Treated/control) | Mechanical Ventilation (Treated/control) | Viral clearance control/ cortico |
|---|---|---|---|---|---|---|---|---|---|---|---|---|---|---|---|---|---|---|
| 1 | Yam et al., 2007 | Corticosteroid Treatment of Severe Acute Respiratory Syndrome in Hong Kong | 2007 | Retrospective study (Inconclusive) | SARS | hydrocortisone; prednisolone; methylprednisolone | 1188/99 | 17%(202/1188)/ 28.3% (28/99) | 4.2%/2% | | 43/57 | 42/58/ 52/48 | >18 yrs/>18 yrs | | | | | |
| 2 | Boudreault et al., 2011 | Impact of Corticosteroid Treatment and Antiviral Therapy on Clinical Outcomes in Hematopoietic Cell Transplant Patients Infected with Influenza Virus | 2011 | Retrospective single-centered study | influenza | prednisone/ methylprednisolone, beclomethasone dipropionate (BDP) | 80/63 | 12.5% (10/80)/ 11.11% (7/63) | 21% (63) Control, 19 (43)% low, 19(37)% high dose | | 83/60 | 58%/42% /57%/43% | 42 (31 - 53) - median/42 | | | 4(10%)/9 (14%) | - | -/7 (5-12) |
| 3 | Moreno et al., 2018 | Corticosteroid Treatment in Critically Ill Patients With Severe Influenza Pneumonia: A Propensity Score Matching Study | 2018 | prospective cohort study | influenza A(H1N1)pdm09 virus | 578 (95.7%) methylprednisolone; 23 (3.8%) prednisolones; 3 (0.5%) dexamethasone | 604/1242 | 27.50%/18.80% | 23%/20% | 7.6% / 6.4 % | 59.37/40.63 | 59.1/40.9/59.5/ 40.5% | 52/ No data | All were ICU patients | 8-Oct | 506 (83.8%)/921 (74.2%) | 8 (3-17)/ 8 (3-16) | |
| 4 | Diaz et al., 2012 | Corticosteroid Therapy in Patients With Primary Viral Pneumonia Due to Pandemic (H1N1) 2009 Influenza | 2012 | Prospective, observational | Influenza (H1N1) | corticosteroid | 136/136 | 18.38%/17.37% | - | - | 55/45 | 57.60%/50.7% (69/67) | 43/ No data | | | - | 9.44 (14.36)/ 9.47 (13.11) | |
| 5 | Martin-Loeches et al., 2011 | Use of early corticosteroid therapy on ICU admission in patients affected by severe pandemic (H1N1)v influenza A infection | 2011 | Prospective, observational | H1N1 Influenza A | corticosteroid | 126/94 | 46.00%/18.10% | 26.2/13.8% | | 113/107 | 54/46/ 47.9/52.1 | 43.26(11.2)/ 46.1 (17.4) | | 12.9+- 14/10.8+-9.8 | | | |
| 6 | Delaney et al., 2016 | The influence of corticosteroid treatment on the outcome of influenza A(H1N1pdm09)-related critical illness | 2016 | observational cohort study | H1N1pdm09 | prednisone, Hydrocortisone and Methylprednisolone, Cortisone, Dexamethasone | 280/327 | 25.50%/16.40% | 87 (31.1%) / 118 (36.1%) | No data | 48.1/51.9 | 45.7/54.3/ 50.1/49.9 | 47.4/ No data | All were ICU patients | 18.5/14.8 | 260 (93.5)/ 275 (85.4) | 15.5 ± 10.1/12.3 ± 10.1 | |
| 7 | Arabi et al., 2018 | Corticosteroid Therapy for Critically ill Patients with the Middle East Respiratory Syndrome (retrospective cohort) | 2018 | retrospective cohort study | MERS-CoV | Hydrocortisone-methylprednisolone, dexamethasone, prednisolone | 151/158 | 74.20%/57.60% | - | - | 213/94 | 70.9/29.1/ 67.1/22.9 | 56.7/ No data | 21.0 (13-38) /15.0 (8-30) | 12.5 (8-23) / 7.0 (5-13) | 93.4% (141/151)/ 76.6% (121/158) | | |
| 8 | Kim et al., 2011 | Corticosteroid Treatment in Critically Ill Patients with Pandemic Influenza A/H1N1 2009 Infection | 2011 | cohort study | Influenza A H1N1 | prednisolone, Hydrocortisone and Methylprednisolone | 107/138 | 58%/27% | 57%/22% | - | 54.69/45.31 | 57/43/ 52.9/47.1 | 55.32/ No data | 30.8 (36.9)/18.9 (20.0) | 13.5 (13.2) /8.8 (9.2) | 85% (91/107)/ 51% (71/138) | 13.3 (13.2)/ 9.6 (10.0) | |
| 9 | Brun-Buisson et al., 2011 | Early Corticosteroids in Severe Influenza A/H1N1 Pneumonia and Acute Respiratory Distress Syndrome | 2011 | retrospective analysis | Influenza A/H1N1 | Hydrocortisone, Methylprednisolone, Prednisone | 83/125 | 33.70%/16.80% | 27.7% /34.4% | | 50.5%/49.5% | 45.4%/56.6%/ 55.2%/44.8% | 47 (35-55)/ 45 (42-56) | | 22/17 | | 17 (10-29)/ 13 (8-24) | |
| 10 | Villar et al., 2020 | Dexamethasone treatment for the acute respiratory distress syndrome: a multicentre, randomised controlled trial | 2020 | randomised controlled trial | None | dexamethasone | 139/138 | 21/36 | 24/25 | | 61/39 | | 57 | | | | 15.7/20.5 | |
| 11 | Li et al., 2017 | Effect of low-to-moderate-dose corticosteroids on mortality of hospitalized adolescents and adults with influenza A(H1N1)pdm09 viral pneumonia | 2017 | Case-Control | Influenza A(H1N1) pdm09 | Methylprednisolone, Dexamethasone, Hydrocortisone, Prednisolone | 1055/1089 | 30 day: 22%, 60-day: 24.7%/ 30 day: 6.8%, 60-day: 7% | 19.1%/4.1% | | 51.1/48.9 | 50.2/49.8/ 52/48 | 34.4/ No data | no data | no data | Invasive mechanical ventilation: 367 (34.8%)/49 (4.5%) | no data | |
| 12 | Cao et al., 2016 | Adjuvant corticosteroid treatment in adults with influenza A (H7N9) viral pneumonia | 2016 | cohort study | Influenza A (H7N9) | Methylprednisolone, Dexamethasone, Hydrocortisone | 65/65 | 27(41.5%)/ 10 (15.3%) | 49.4/41.6 | - | 70/30 | 70.8/29.2/ 69.2/30.8 | 58 (46–65) + 56 (43–68) | - | - | 38/27 | - | |
| 13 | Linko et al., 2011 | Corticosteroid therapy in intensive care unit patients with PCR-confirmed influenza A(H1N1) infection in Finland | 2011 | prospective observational study | Influenza A(H1N1) | methylprednisolone, Hydrocortisone | 72/60 | 11%/2% | 55.9/12.8% | n/a | n/a | 64/36/65/35 | n/a | 24 [14–37]/ 15 [8–25] | 13 [18–20]/4 [3–5] | 50.70%/34.80% | 6 (1-12)/0 | |
| 14 | Lu et al., 2020 | Adjuvant corticosteroid therapy for critically ill patients with COVID-19 | 2020 | retrospective cohort study | SARS Cov2 | methylprednisolone 1:5, dexamethasone 1:25) | 151/93 | 52.30%/5.40% | | | 52/48 | 55/45/48/52 | 62 (50-71) | | | 78 (52%)/4 (4%) | | |
| 15 | Hong et al., 2020 | Corticosteroid treatment in patients with severe covid-19 pneumonia | 2020 | Retrospective cohort study | SARS-CoV-2 | oral prednisone and systemic methylprednisolone. | 93/84 | 53%/57% | - | - | - | - | - | - | - | 38/93/10/84 | - | |
| 16 | Liu et al., 2020 | Low-to-moderate dose corticosteroids treatment in hospitalized adults with COVID-19 | 2020 | Retrospective cohort study | SARS-CoV-2 | Methylprednisolone, Dexamethasone, Hydrocortisone, Prednisolone | 124/124 | 37.9/37.9% | 8.9%/5.6% | | | | | 44147 | 44020 | | | |
| 17 | Fernández-Cruz et al., 2020 | A Retrospective Controlled Cohort Study of the Impact of Glucocorticoid Treatment in SARS-CoV-2 Infection Mortality | 2020 | Retrospective Controlled Cohort Study | SARS-CoV-2 | methylprednisolone | 396/67 | 55/396/16/67 | - | - | - | 276/120/41/16 | 65.4 | - | - | - | - | |
| 18 | Li et al., 2020 | Efficacy Evaluation of Early, Low-Dose, Short-Term Corticosteroids in Adults Hospitalized with Non-Severe COVID-19 Pneumonia: A Retrospective Cohort Study | 2020 | retrospective cohort study | SARS-CoV-2 | | 55/55 | 1.89/0% | 89.1 | 23.6 | | | | 23/15 | | | | 11/18 day |
| 19 | Ma et al., 2020 | Corticosteroid therapy for patients with severe novel Coronavirus disease 2019 | 2020 | retrospective cohort study | SARS-CoV-2 | | 47/25 | 4.2/8% | | 31/18 | | | | 18.7/21 | | 24/8 | 9.6/12.8 | 19.4/16.1 |
| 20 | Dequin et al., 2020 | Effect of Hydrocortisone on 21-Day Mortality or Respiratory Support Among Critically Ill Patients With COVID-19 | 2020 | RCT | SARS-CoV-2 | hydrocortisone | 76/73 | 14.7/27.4% | | 37.3%/41.1% | 69.8/30.2 | | 62.2 | | | 0.227/0.233 | | |
| 21 | Tomazini et al., 2020 | Effect of Dexamethasone on Days Alive and Ventilator-Free in Patients With Moderate or Severe Acute Respiratory Distress Syndrome and COVID-19: The CoDEX Randomized Clinical Trial | 2020 | RCT | SARS-CoV-2 | dexamethasone | 151/148 | 56.3/61.5% | 7.9/9.5 | 21.9/29.1 | | | | | | | 12.5/13.9 | |
| 22 | Horby et al., 2020 | Dexamethasone in Hospitalized Patients with Covid-19 — Preliminary Report RECOVERY TRIAL | 2020 | RCT | SARS-CoV-2 | dexamethasone | 2104/4321 | 22.9/25.7% | | | | | | | | 228/400 | | |
| 23 | Angus et al., 2020 | Effect of Hydrocortisone on Mortality and Organ Support in Patients With Severe COVID-19 The REMAP-CAP COVID-19 Corticosteroid Domain Randomized Clinical Trial | 2020 | RCT | SARS-CoV-2 | hydrocortisone | 278/101 | 28.05/33% | | | | | | | | | | |
| 24 | Jeronimo et al., 2020 | Methylprednisolone as Adjunctive Therapy for Patients Hospitalized With COVID-19 (Metcovid): A Randomised, Double-Blind, Phase IIb, Placebo-Controlled Trial | 2020 | RCT | SARS-CoV-2 | Methylprednisolone | 194/199 | 37.11%/38.19% | | | 64.9/35.1 | 64.3/35.7/ 64.3/35.7 | 55 | 10 (7-13)/9 (7-12) | | 66/194/67/199 | | |

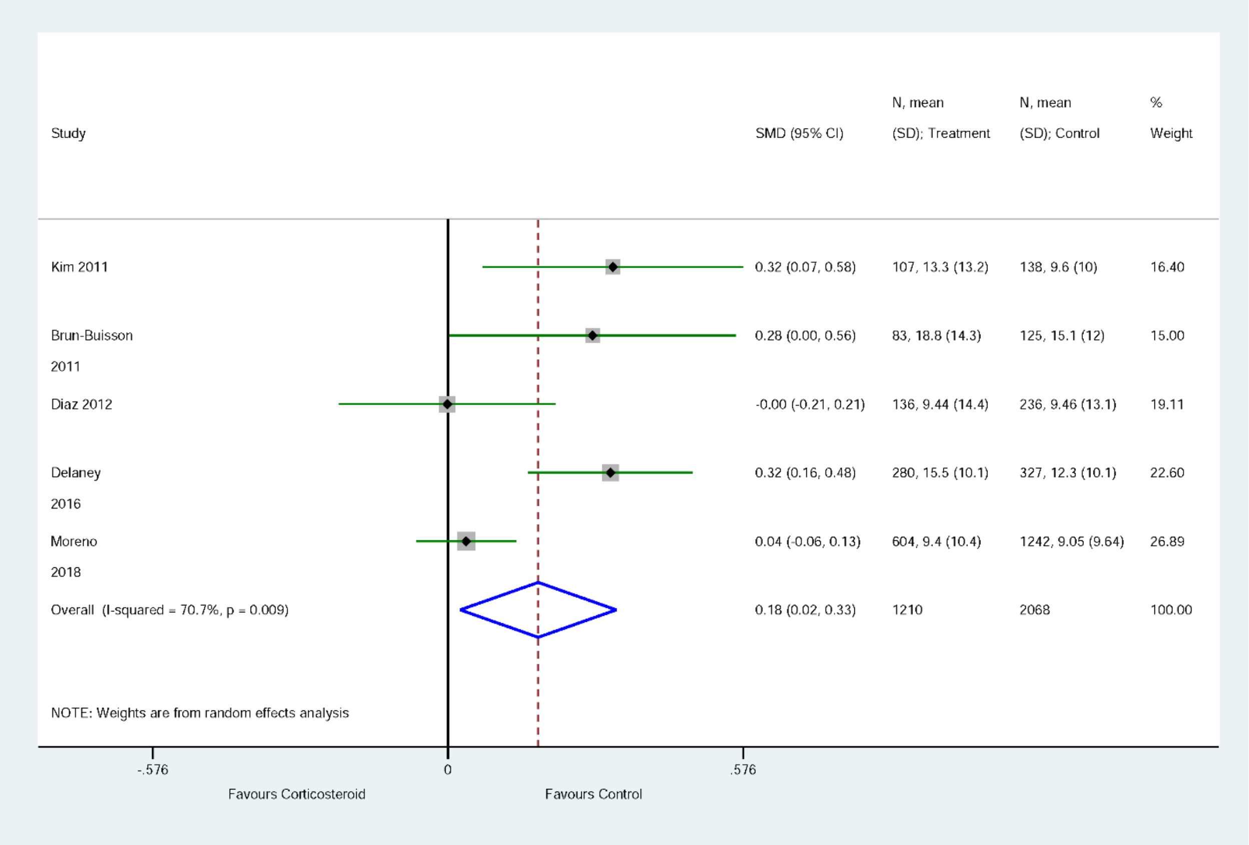


**Suppl. figure S1**. Effect of corticosteroid on Length of Mechanical Ventilation in days (Z=2.26, P=0.024, SMD: standard mean difference, SD: standard deviation, horizontal line express 95% CI, Diamond represents overall estimate from the meta-analysis, squares represent effect size for each study).

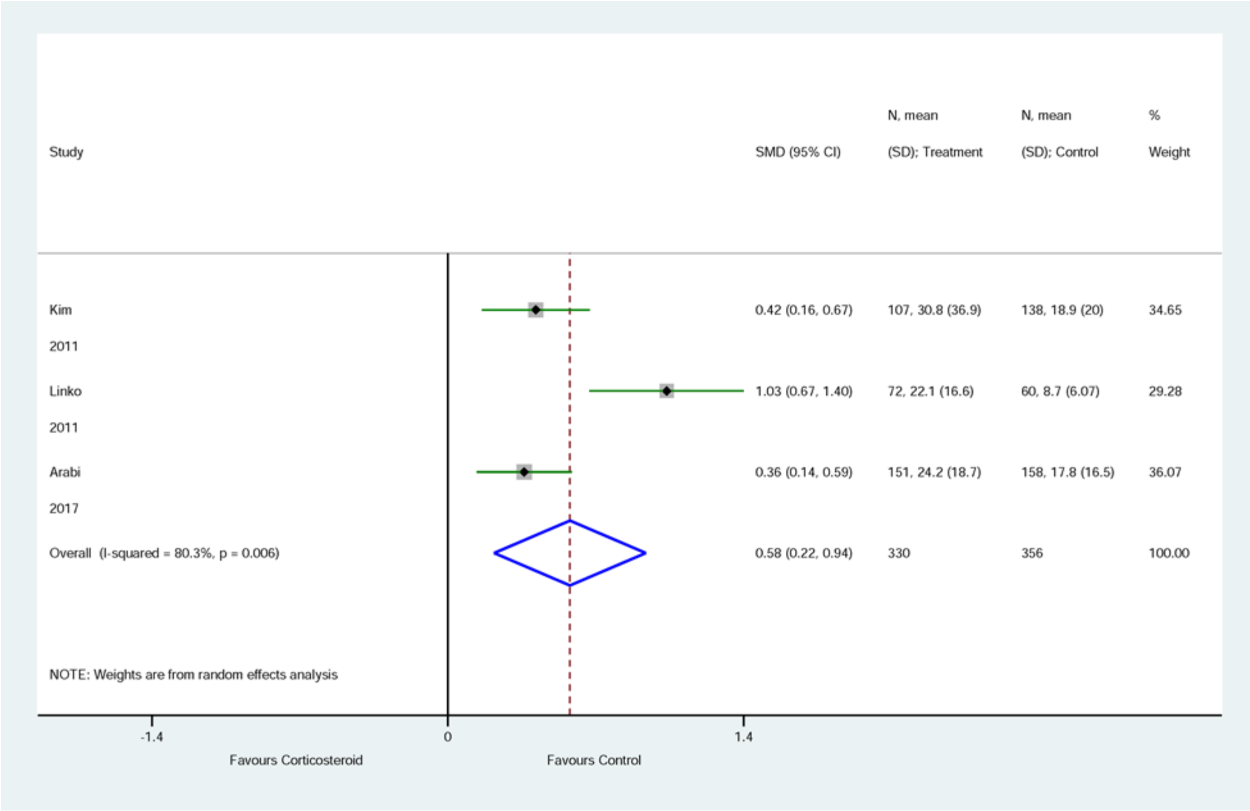


**Suppl. figure S2**. Effect of corticosteroid on the length of hospital stay (Z=3.16, P=0.002, SMD: standard mean difference, SD: standard deviation, horizontal line express 95% CI, Diamond represents overall estimate from the meta-analysis, squares represent effect size for each study).

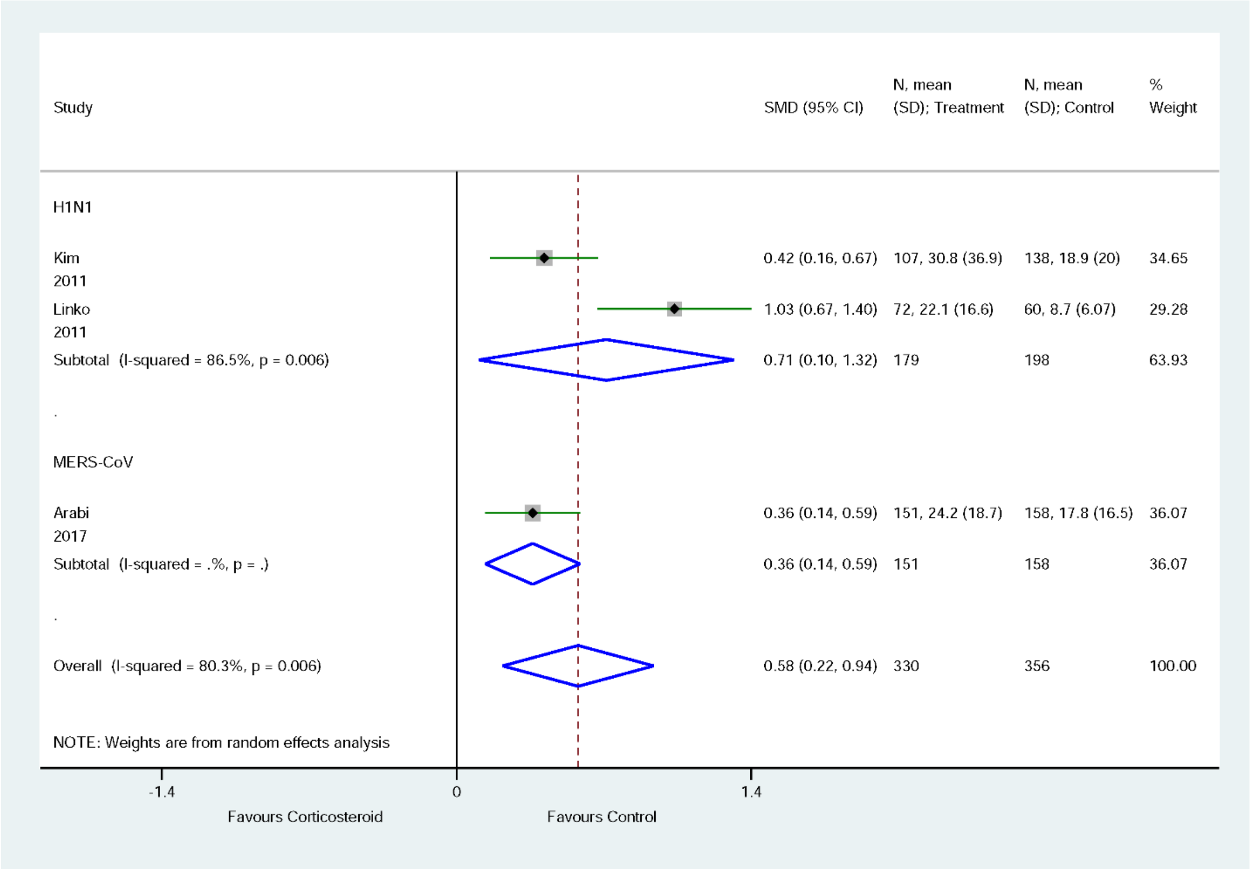


**Suppl. figure S3**. Subgroup analysis of effect of corticosteroid on length of hospital stay for patients with H1N1 and MERS-CoV viral infections (SMD: standard mean difference, SD: standard deviation, horizontal line express 95% CI, Diamond represent overall estimate from the meta-analysis, squares represent effect size for each study).

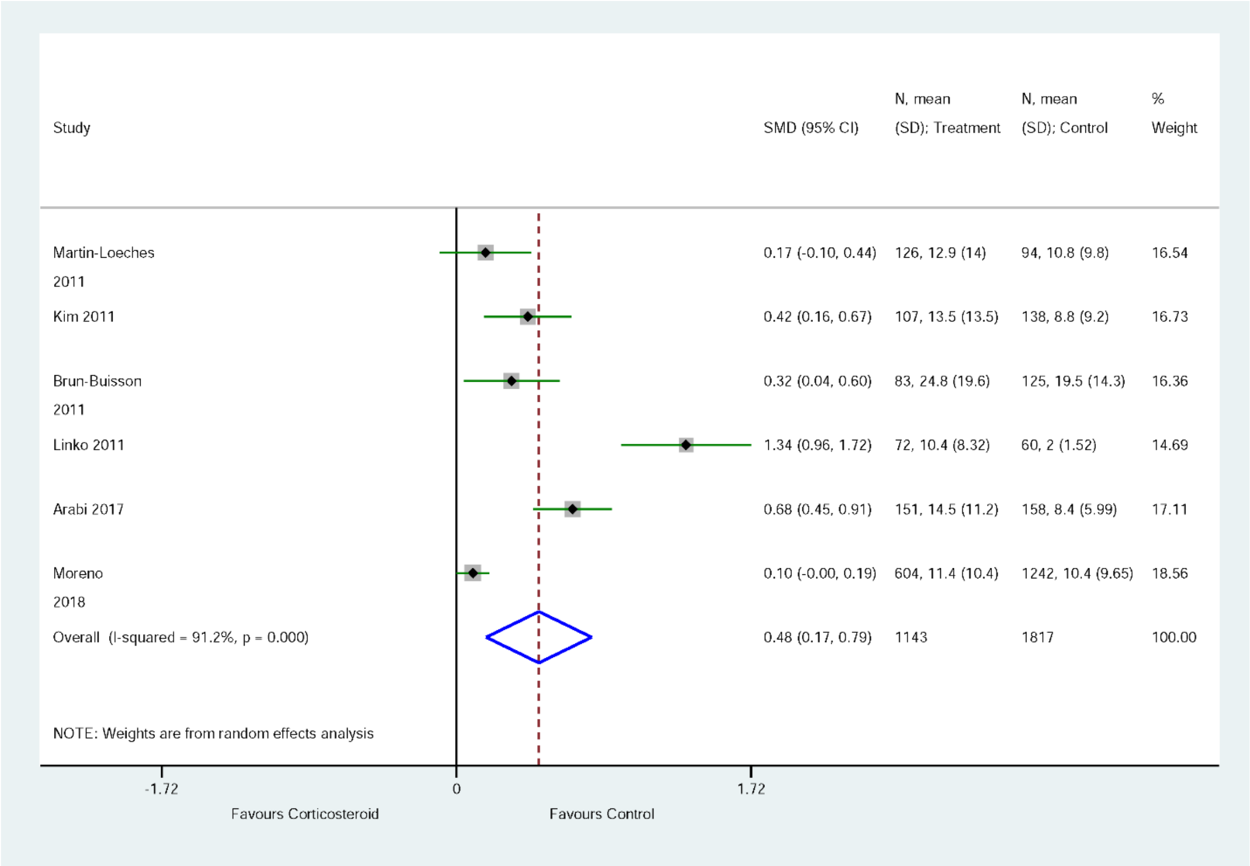


**Suppl. figure S4**. Effect of corticosteroid on the length of ICU stay (Z=3.07, P=0.002, SMD: standard mean difference, SD: standard deviation, horizontal line express 95% CI, Diamond represents overall estimate from the meta-analysis, squares represent effect size for each study).

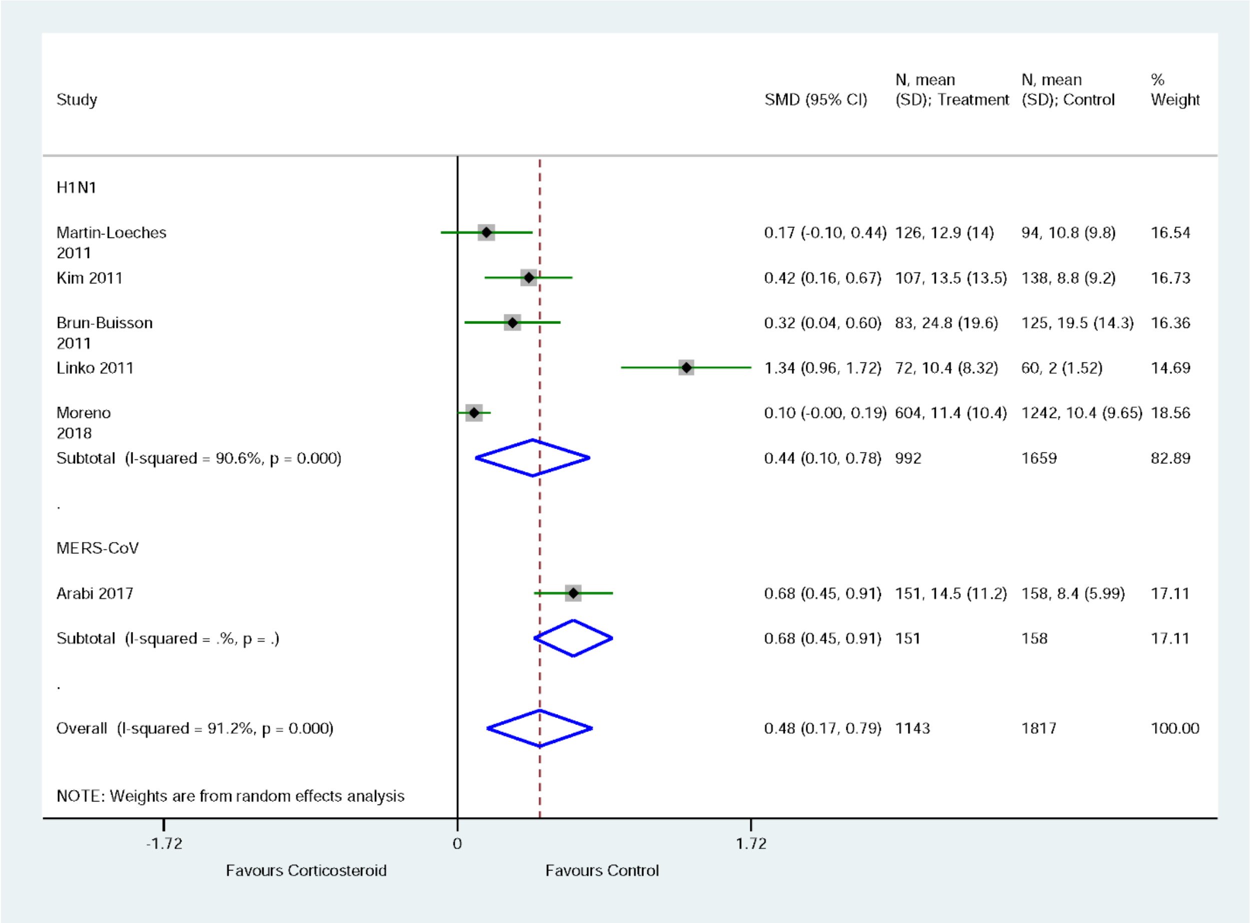


**Suppl. figure S5.** Subgroup analysis of effect of corticosteroid on length of ICU stay for patients with H1N1 and MERS-CoV viral infection (SMD: standard mean difference, SD: standard deviation, horizontal line express 95% CI, Diamond represent overall estimate from the meta-analysis, squares represent effect size for each study).

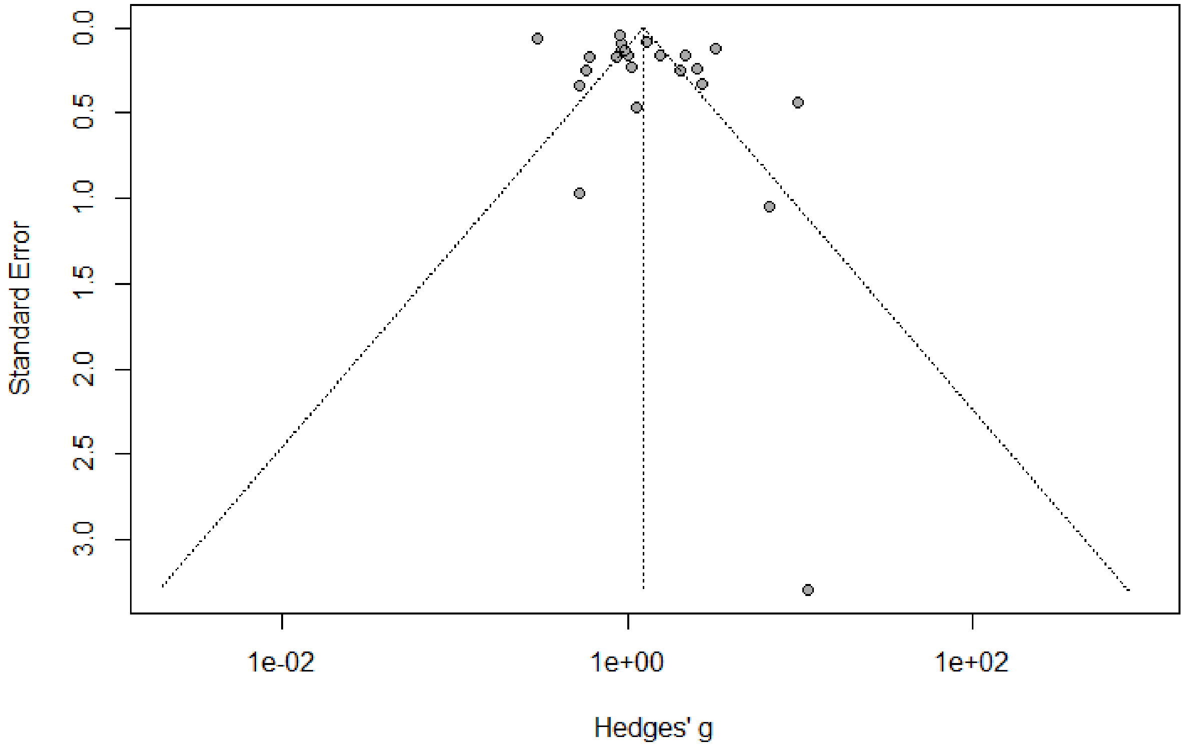


**Suppl. figure S6**. Funnel plot analysis of selected articles to check publication bias.

**Egger's test (Egger, Smith et al. 1997):**

Intercept: 2.788
Confidence interval: - -0.348 to -5.924
t statistic: 1.766
P value: 0.09

**Interpretation:**

We can see that the P value of Egger's test is not significant (P > 0.05), which suggest that there is no evidence of publication bias.

Egger, M., et al. (1997). "Bias in meta-analysis detected by a simple, graphical test." Bmj **315**(7109): 629-634.

**Suppl. Table ST3**: Risk of bias of included Observational study

| Study | A. Selection | | | | B. Comparability of cohort | C. Outcome | | |
|---|---|---|---|---|---|---|---|---|
| | Represent-activeness of exposed cohort | Selection of non-exposure | Ascertainment of exposure | Outcome not present at start | | Assessment of exposure | Follow-up long enough? | Adequacy of Follow-up |
| Yam 2007 | ☆ | ☆ | ☆ | ☆ | | ☆ | ☆ | ☆ |
| Boudreault 20111 | ☆ | ☆ | ☆ | ☆ | | ☆ | | ☆ |
| Moreno 2018 | ☆ | ☆ | ☆ | ☆ | ☆☆ | ☆ | | ☆ |
| Diaz 2012 | ☆ | ☆ | ☆ | ☆ | | ☆ | ☆ | ☆ |
| Martin-Loeches 2018 | ☆ | ☆ | ☆ | ☆ | | ☆ | | ☆ |
| Delaney 2016 | ☆ | ☆ | ☆ | ☆ | | ☆ | | ☆ |
| Lu 2020 | ☆ | ☆ | ☆ | ☆ | ☆☆ | ☆ | | ☆ |
| Arabi 2018 | ☆ | ☆ | ☆ | ☆ | | ☆ | ☆ | ☆ |
| Kim 2011 | ☆ | ☆ | ☆ | ☆ | ☆☆ | ☆ | ☆ | ☆ |
| Brun-Buisson 2011 | ☆ | ☆ | ☆ | ☆ | ☆☆ | ☆ | | ☆ |
| Li 2017 | ☆ | ☆ | ☆ | ☆ | ☆☆ | ☆ | ☆ | ☆ |
| Cao 2016 | ☆ | ☆ | ☆ | ☆ | ☆☆ | ☆ | ☆ | ☆ |
| Linko 2011 | ☆ | ☆ | ☆ | ☆ | | ☆ | | ☆ |
| Hong et al., 2020 | ☆ | ☆ | ☆ | ☆ | ☆ | ☆ | ☆ | ☆ |
| Liu et al., 2020 | ☆ | ☆ | ☆ | ☆ | ☆☆ | ☆ | | ☆ |
| Fernández-Cruz et al., 2020 | ☆ | ☆ | ☆ | ☆ | | ☆ | ☆ | ☆ |
| Li et al., 2020 | ☆ | ☆ | ☆ | ☆ | ☆☆ | ☆ | ☆ | ☆ |
| Ma et al., 2020 | ☆ | ☆ | ☆ | ☆ | | ☆ | | ☆ |

Stars indicate the scores assigned to each study

**Suppl. Table ST4**: Risk of bias of included randomized controlled trials studies

| Study | Sequence Generation | Allocation Sequence Concealment | Blinding (Performance bias) | Blinding (Outcome measurement) | Missing Outcome Data | Other Bias | Overall Bias |
|---|---|---|---|---|---|---|---|
| Villar 2020 | Low | Low | Low | Low | Low | Probably Low | Low |
| Dequin et al., 2020 | Low | Low | Low | Low | Low | Probably Low | Low |
| Tomazini et al., 2020 | Low | Low | Low | Low | Low | Probably Low | Low |
| Horby et al., 2020 | Low | Low | Low | Low | Low | Probably Low | Low |
| Angus et al., 2020 | Low | Low | Low | Low | Low | Probably Low | Low |
| Jeronimo et al., 2020 | Low | Low | Low | Low | Low | Probably Low | Low |
